# SCOPE: AI-Assisted Early Detection of Potentially Curable Pancreatic Neoplasms on CT from Local and Global Information

**DOI:** 10.64898/2026.02.04.26345495

**Authors:** Felipe Oviedo, Felipe Lopez-Ramirez, Alejandra Blanco, James Facciola, Stephen Kwak, Jason M. Zhao, Emir A. Syailendra, Florent Tixier, Rahul Dodhia, Ralph H. Hruban, William B. Weeks, Juan M. Lavista Ferres, Linda C. Chu, Elliot K. Fishman

## Abstract

**Purpose:** To develop SCOPE (Small-lesion COntextual Pancreatic Evaluator), a deep learning model designed to improve CT detection of small pancreatic lesions—pancreatic ductal adenocarcinoma (PDAC), pancreatic neuroendocrine tumors (PanNETs), and cystic lesions—by integrating voxel-level features with global context.

**Materials and Methods:** This retrospective study used three independent datasets. A development cohort of 4,065 contrast-enhanced CT scans was used to train a deep neural network that performs pancreas, ductal, and lesion segmentation with an integrated classification head. A metamodel combined segmentation-derived and global contextual signals for case-level prediction. Performance was assessed on (1) an internal holdout test set (*n* = 605), (2) an external multi-institutional PDAC dataset from the PANORAMA challenge (*n* = 2,238), and (3) an expert-curated small-lesion reader study (*n* = 200). Areas under the receiver operating characteristic curve (AUCs) were compared using DeLong test; sensitivities and specificities using McNemar’s test.

**Results:** On the internal test set, SCOPE improved lesion-versus-normal AUC compared with the best segmentation baseline (0.974 [95% CI: 0.964, 0.984] vs 0.956; *P* = .006) and increased small-lesion sensitivity at 95% specificity (0.727 [95% CI: 0.653, 0.801] vs 0.600; *P* = .012). Performance gains were observed across lesion classes, with significant improvements for PDAC and PanNET detection. On the external dataset, SCOPE improved PDAC-versus-non-PDAC AUC (0.978 vs 0.861, *P* < .001) and achieved higher sensitivity at 90% and 95% specificity without retraining. For the small-lesion reader study, SCOPE achieved lesion-versus-normal AUC of 0.922 and performed within the range of subspecialty abdominal radiologists; SCOPE provided the correct diagnosis in 14.5% (29/200) of cases in which two or more readers were incorrect.

**Conclusion:** SCOPE improves early detection of small, potentially curable, pancreatic lesions on CT by combining local segmentation and global pancreatic context. Its consistent performance across internal, external, and reader datasets supports potential use as a concurrent reader for earlier and more accurate pancreatic lesion detection.

## 1 Introduction

Pancreatic cancer is projected to become the second leading cause of cancer death worldwide, reflecting both rising incidence and the fact that most patients present with advanced disease (1). Pancreatic ductal adenocarcinoma (PDAC) remains one of the deadliest pancreas malignancies, with 5-year survival typically below 15%. Survival outcomes improve substantially for patients with carcinomas ≤ 2 cm, where reported 5-year survival rates reach 30–60% (2,3). The sensitivity of radiologists for detecting small PDACs on computed tomography (CT) can be as low as 60% (4) because these small lesions often do not form a discrete, clearly visible mass, and instead manifest through indirect findings such as focal pancreatic duct cutoff, upstream ductal dilation, or parenchymal atrophy (3,5). Similar size-dependent considerations apply to pancreatic neuroendocrine tumors (PanNETs) and small pancreatic cystic lesions, e.g., intraductal papillary mucinous neoplasms (IPMNs) and mucinous cystic neoplasms (MCNs), where sub-2 cm lesions have the potential to progress to invasive cancer and management depends on early recognition of subtle ductal or morphologic changes (6–8).

Deep learning approaches have recently achieved high performance in the detection of pancreatic tumors, particularly for PDAC, in some settings matching or exceeding average performance of radiologists (9–13). However, the relevance of these deep learning approaches to the detection of small, potentially curable, lesions is limited for three main reasons. First, most methods have been trained using healthy controls, simplifying the task and failing to capture the real distribution of pancreatic findings, where cystic lesions, PanNETs, and normal variants frequently co-exist and must be distinguished from early malignancy. Second, performance on small lesions—the subgroup most relevant for successful intervention—is rarely reported explicitly, and when reported, detection performance decreases markedly (3,9,14). Third, most deep learning pipelines that incorporate secondary signs or ductal morphology still rely on localized patches or engineered features without modeling whole-gland context (3,15–19). Beyond PDAC, automated detection of small PanNETs and cystic lesions remains limited. Although recent work shows improved performance versus normal controls (10,16,20) or promising results for scalable annotation (12), existing methods have not been evaluated in the more challenging setting of detecting small lesions across multiple pancreatic lesion types, where subtle secondary signs and overlapping imaging patterns limit performance.

We hypothesized that an end-to-end model integrating local voxel-level features with global pancreatic context would improve the detection of small, potentially curable, lesions. In this study, we introduce SCOPE (Small-Lesion COntextual Pancreatic Evaluator), a deep learning model for detecting small pancreatic lesions across PDAC, PanNET, and cystic subtypes. SCOPE integrates pancreas, duct, and lesion segmentation with a classification module that captures gland morphology and secondary imaging features. We trained and evaluated SCOPE on one of the largest contrast-enhanced CT datasets of its kind.

## 2 Materials and Methods

### 2.1 Study design and datasets

This retrospective study was HIPAA-compliant and IRB-approved. Three datasets were used (Figure 1A), all contrast-enhanced venous-phase CT.

**Figure 1:**
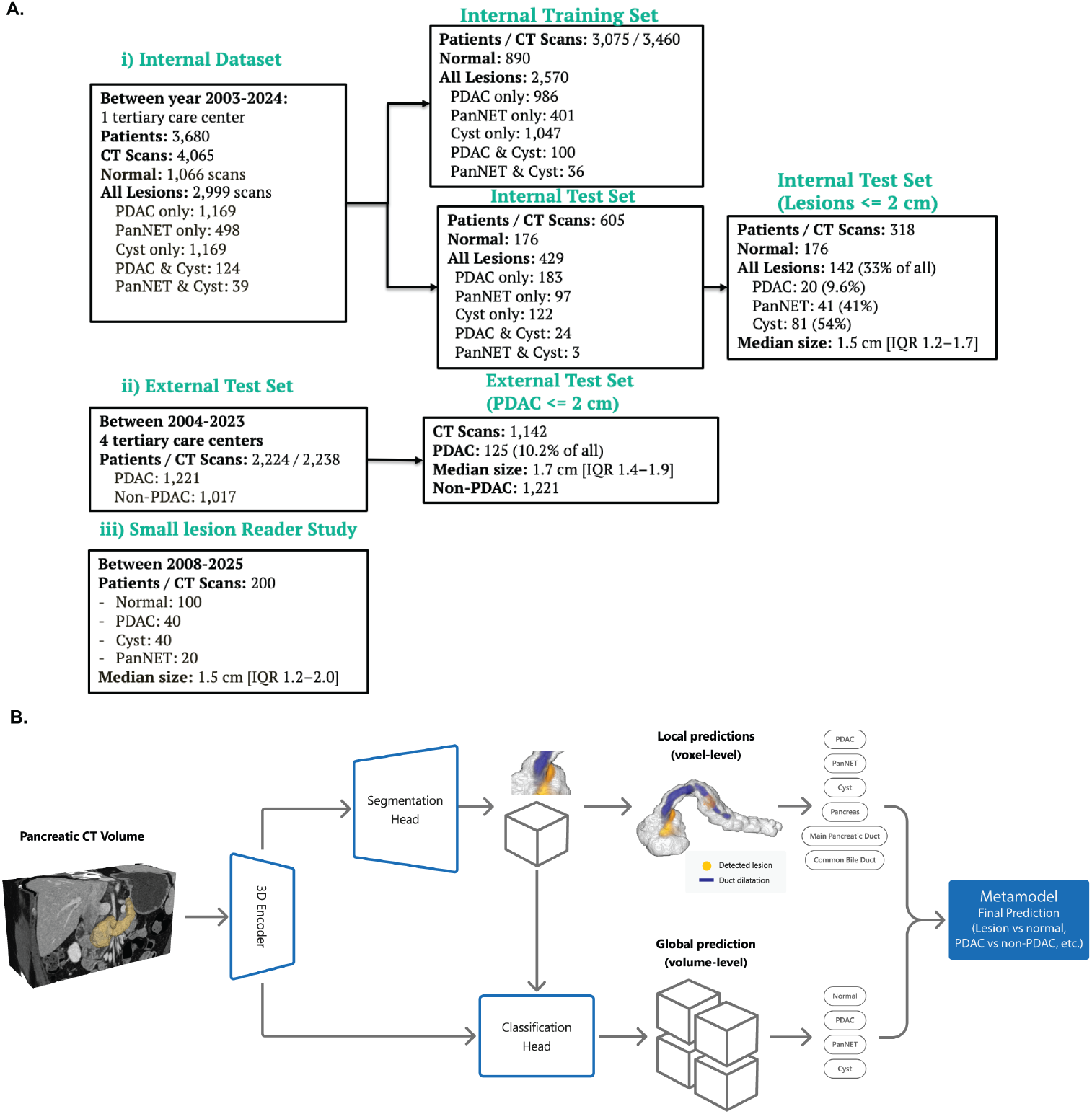
**A)** Contrast-enhanced venous CT datasets used in this study, along with lesion types and proportion of small lesions (*<* 2 cm) in them. Three datasets are considered: (i) internal dataset, external dataset (PANORAMA training dataset), (iii) small lesion reader study. **B)** SCOPE model architecture composed of a 3D encoder, a voxel-level segmentation head, and a case-level classification head. Classes predicted by each head are presented.

#### i) Internal development dataset

4,065 scans from 3,680 patients with PDAC, PanNET, pancreatic cysts, or normal pancreas verified by pathology or institutional tumor board records (Appendix S1). Radiologists performed slice-by-slice segmentation of the pancreas, main pancreatic duct, common bile duct, and lesions. For cysts, although not all were pathologically proven, most were morphologically presumed to be IPMNs, with additional cyst types in Table 1. We split this dataset into training (*n* = 3,460) and holdout test (*n* = 605) sets with no patient overlap; 5.5% of abnormal scans (163/2,964) contained co-occurring cysts and PDAC or PanNET.

**Table 1:** Patient characteristics in the internal test set and in the small lesion reader study cohort.

| Characteristics | Internal Test Set | Small Lesion Reader Study |
| --- | --- | --- |
| No. Patients | 605 | 200 |
| No. CT scans | 605 | 200 |
| Age at time of scan (mean ± SD) | 58.1 ± 14.5 (14 patients not known) | 61.2 ± 10.5 |
| **Sex** |  |  |
| Female | 329 (54.3) | 114 (57.0) |
| Male | 262 (43.3) | 85 (43.0) |
| Not specified | 14 (2.3) | 0 (0.0) |
| **Race / Ethnicity** |  |  |
| American Indian/Alaska Native | 1 | 0 |
| Asian | 19 | 10 |
| Black or African American | 68 | 25 |
| White | 467 | 153 |
| Latino/Hispanic | 15 | 4 |
| Unknown/Other | 34 | 8 |
| **Path Report** |  |  |
| PDAC | 207 | 40 |
| PanNET | 100 | 20 |
| Normal | 176 | 100 |
| Cyst | 149 | 40 |
| Cyst, non-specified | 88 | 8 |
| True cyst (benign epithelial) | 25 | 1 |
| IPMN | 21 | 22 |
| MCN | 10 | 4 |
| SCA | 3 | 3 |
| Other | 2 | 2 |

#### ii) External multi-institutional dataset

2,238 scans from the PANORAMA challenge development cohort (1,221 PDAC; 1,017 non-PDAC cases) (11).

#### iii) Small-lesion reader cohort

200 diagnostically challenging scans (100 lesions ≤ 2 cm, 100 normals) curated by a radiologist with *>* 30 years of experience. Three abdominal radiologists (R1: 6 years post-fellowship, R2: 2 years, R3: 4 years) independently reviewed the venous-phase scan and assigned a mutually exclusive diagnosis (PDAC, PanNET, cyst, or normal).

In datasets (i) and (ii), small lesions were defined as those with a maximal axial connected component diameter of ≤ 2 cm based on ground-truth segmentation; in dataset (iii), small lesions were defined according to the radiologist report and reconfirmed during curation. Patient characteristics for the internal test set and reader cohort are summarized in Table 1.

### 2.2 Model

We developed SCOPE, a multi-head deep learning model composed of a 3D encoder, a 3D segmentation head, and a case-level classification head (Figure 1B). Following pancreas localization and CT cropping using PanSegNet (21), the encoder (MedNeXt (22) or Swin-UNETR (23), Appendix S2) extracted hierarchical volumetric features. The segmentation head generated voxel-level predictions for the pancreas, main pancreatic duct, common bile duct, and pancreatic lesions (PDAC, PanNET, and pancreatic cysts) in a region of interest (ROI). In parallel, the classification head integrated deep encoder features and segmentation information, using 3D coordinate attention to capture gland, lesion, and ductal morphology, identifying secondary signs associated with early disease (5) (e.g., abrupt duct cutoff, ductal dilation, or mild gland atrophy). Because multiple lesion types can coexist, the classification head produced independent sigmoid probabilities for global detection of PDAC, PanNET, and cysts. Case-level predictions were obtained by max-pooling outputs across overlapping ROIs.

Final diagnostic outputs were generated using a lightweight metamodel (FLAML) (24) that integrated global classification probabilities with local information from pooled segmentation masks. By specializing the metamodel for a given task, SCOPE could be adapted to diverse clinical applications (e.g., PDAC vs. non-PDAC classification or triaging any lesion) without retraining the neural network.

Model training was performed in three stages. First, the SCOPE encoder and segmentation head were trained for voxel-level segmentation using Dice–cross-entropy (DiceCE) loss. Second, the classification head was trained using case labels with the encoder and segmentation head frozen, producing global lesion probabilities optimized with binary cross-entropy loss applied independently to each lesion class. Finally, all components were jointly fine-tuned using a weighted combination of voxel-level and case-level losses, while preventing classification gradients from degrading segmentation performance. The model will be available at the time of publication in GitHub. Additional details are provided in Appendix S2.

### 2.3 Model evaluation and baselines

Model performance was evaluated in each test set without retraining using receiver operating characteristic (ROC) curves and area under the curve (AUC). As radiologists typically yield 90–95% specificity when detecting pancreatic lesions of a single class (9), we report model performance at these operating points. Superiority and non-inferiority of AUCs were assessed using DeLong’s test, while sensitivities and specificities were compared using McNemar’s test, with significance level *α* = 0.05 in both cases. Bootstrap (*n* = 10,000) was used to compute confidence intervals.

In the internal and external test sets, segmentation-only models, including MedNeXt (22), SwinUNETR (23), and nnU-Net (25), were trained from scratch on the internal training set using DiceCE loss. We chose the best performing model in both test sets (MedNeXt-M) as the baseline comparison model (Appendix S2). The pre-trained PANORAMA winner (PanDx) (26) was used for comparing PDAC vs. non-PDAC performance in the internal test set. In the small-lesion reader study, SCOPE performance was compared with each individual reader using McNemar’s test. All statistical tests were performed using `statsmodels` 0.14.5.

## 3 Results

### 3.1 Internal test set

On the internal test set, SCOPE demonstrated consistently higher lesion detection performance than the baseline segmentation-only model (Figure 2), and SwinUNETR was the best backbone (Table S1). For lesions of any size versus normal (Figure 2A), SCOPE achieved an AUC of 0.974 (95% CI: 0.964, 0.984), significantly higher than MedNeXt (0.956, *P* = .006). When evaluating each lesion class versus the rest (Figure 2B), SCOPE yielded higher AUCs for PDAC (0.968 [95% CI: 0.955, 0.982] vs 0.949; *P* = .004) and PanNET (0.950 [95% CI: 0.924, 0.975] vs 0.884; *P* = .002), while cyst detection remained similar between models (*P* = .288).

**Figure 2:**
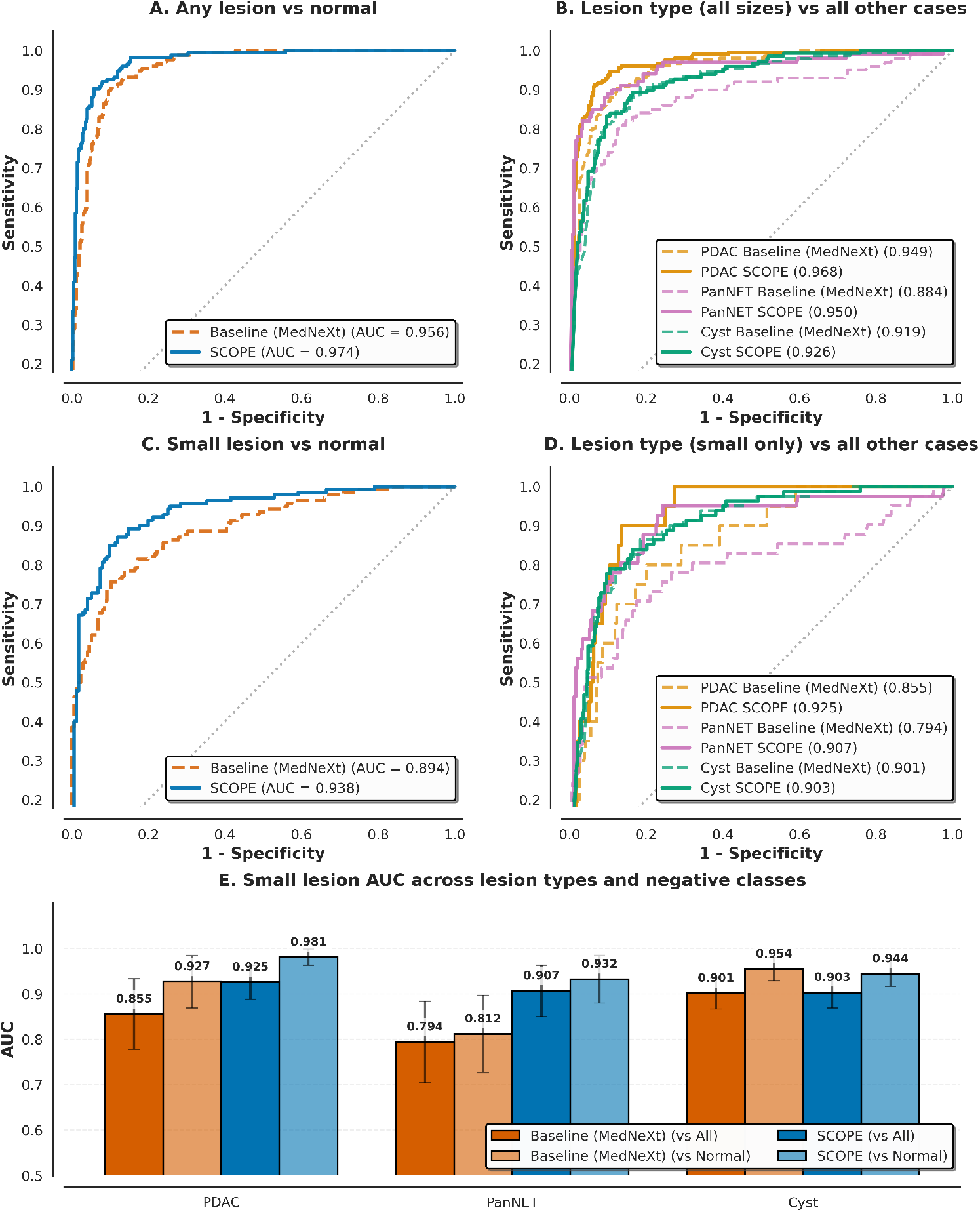
SCOPE performance in the internal test set. **A), B)** Performance for detection of any lesion vs normal, and a specific lesion class vs all others (including normal controls and other lesion types). **C) and D)** summarize performance for small lesions (≤ 2 cm) in the same conditions. **E)** Small lesion AUC across lesion types, illustrating the effect of negative class definition on detection performance. Error bars are 95% confidence inte7rvals of the mean.

Performance gains were most pronounced for small lesions. For small lesions of any type (Figure 2C, Table 2), SCOPE achieved an AUC of 0.938 (95% CI: 0.912, 0.964), exceeding MedNeXt (0.894, *P* = .005). We evaluated two negative-class formulations: (i) a heterogeneous versus-all definition, in which each lesion class of dimension ≤ 2 cm was discriminated from both normal controls and lesions of all sizes of different class, and (ii) a versus-normal definition, consisting only of normal pancreas. Under the more challenging versus-all formulation (Figure 2D), SCOPE outperformed MedNeXt for small PDAC (0.925 [95% CI: 0.888, 0.963] vs 0.855; *P* = .004) and PanNET (0.907 [95% CI: 0.850, 0.961] vs 0.794; *P* = .007), while cyst performance remained comparable. Restricting the negative class to normals only (Figure 2E), AUCs increased for all lesion types; SCOPE again significantly outperformed MedNeXt for PDAC (0.981 [95% CI: 0.963, 0.998] vs 0.927; *P* = .012) and PanNET (0.932 [95% CI: 0.879, 0.985] vs 0.812; *P* = .004), while cyst AUCs remained similar.

**Table 2:** SCOPE, Baseline and PANORAMA winner (PanDx) evaluation results on the internal test set. All lesions are ≤ 2 cm. Best models which are statistically significant are in **bold**. P-values in Appendix S3, Table S4. 95% confidence intervals of mean reported in brackets.

| Metric | Target | SCOPE |  | Baseline (MedNeXt) |  | PANORAMA winner (PanDx) |  |
| --- | --- | --- | --- | --- | --- | --- | --- |
|  |  | vs Normal | vs All | vs Normal | vs All | vs Normal | vs All |
| AUC | Lesion | **0.938**<br>[0.912, 0.964] | – | 0.894<br>[0.859, 0.930] | – | 0.723<br>[0.664, 0.782] | – |
|  | PDAC | **0.981**<br>[0.963, 0.998] | **0.925**<br>[0.888, 0.963] | 0.927<br>[0.869, 0.985] | 0.855<br>[0.777, 0.933] | **0.994**<br>[0.986, 1.000] | **0.921**<br>[0.890, 0.952] |
|  | PanNET | **0.932**<br>[0.879, 0.985] | **0.907**<br>[0.850, 0.963] | 0.812<br>[0.726, 0.897] | 0.794<br>[0.704, 0.884] | – | – |
|  | Cyst | **0.944**<br>[0.916, 0.972] | **0.903**<br>[0.869, 0.937] | **0.954**<br>[0.929, 0.980] | 0.901<br>[0.867, 0.935] | – | – |
| Sensitivity @ 90% Specificity | Lesion | **0.850**<br>[0.791, 0.909] | – | 0.743<br>[0.670, 0.815] | – | 0.450<br>[0.368, 0.532] | – |
|  | PDAC | **0.950**<br>[0.854, 1.000] | **0.750**<br>[0.611, 0.889] | 0.812<br>[0.641, 0.983] | 0.600<br>[0.442, 0.758] | 0.654<br>[0.446, 0.863] | **0.615**<br>[0.459, 0.772] |
|  | PanNET | **0.815**<br>[0.697, 0.934] | **0.734**<br>[0.634, 0.833] | 0.549<br>[0.397, 0.702] | 0.537<br>[0.424, 0.649] | – | – |
|  | Cyst | 0.835<br>[0.754, 0.915] | **0.778**<br>[0.711, 0.844] | **0.877**<br>[0.805, 0.948] | **0.728**<br>[0.657, 0.800] | – | – |
| Sensitivity @ 95% Specificity | Lesion | **0.727**<br>[0.653, 0.801] | – | 0.600<br>[0.519, 0.681] | – | 0.414<br>[0.333, 0.496] | – |
|  | PDAC | **0.900**<br>[0.769, 1.000] | **0.400**<br>[0.242, 0.558] | 0.700<br>[0.499, 0.901] | **0.350**<br>[0.196, 0.504] | 0.624<br>[0.411, 0.836] | **0.350**<br>[0.198, 0.504] |
|  | PanNET | **0.683**<br>[0.540, 0.825] | **0.632**<br>[0.524, 0.741] | 0.512<br>[0.359, 0.665] | 0.491<br>[0.378, 0.603] | – | – |
|  | Cyst | 0.751<br>[0.656, 0.845] | **0.595**<br>[0.516, 0.673] | **0.814**<br>[0.729, 0.898] | **0.528**<br>[0.449, 0.608] | – | – |

At high-specificity operating points (Figure 3A, Table 2), SCOPE achieved higher lesion sensitivity than the segmentation-only baseline on the internal test set. At 90% specificity, lesion sensitivity increased from 0.743 to 0.850 (95% CI: 0.791, 0.909), *P* = .047 for SCOPE, and at 95% specificity, sensitivity improved from 0.600 to 0.727 (95% CI: 0.653, 0.801), *P* = .006. In the PDAC-vs-normal setting, SCOPE achieved sensitivity of 0.950 (95% CI: 0.854, 1.00) at 90% and 0.900 (95% CI: 0.769, 1.00) at 95% specificity, exceeding MedNeXt (0.812 and 0.700, *P* < .001 for both). Mean dice scores of SCOPE predictions were 0.611 (95% CI: 0.583, 0.639) for all lesions and 0.493 (95% CI: 0.437, 0.548) (Table S2).

**Figure 3:**
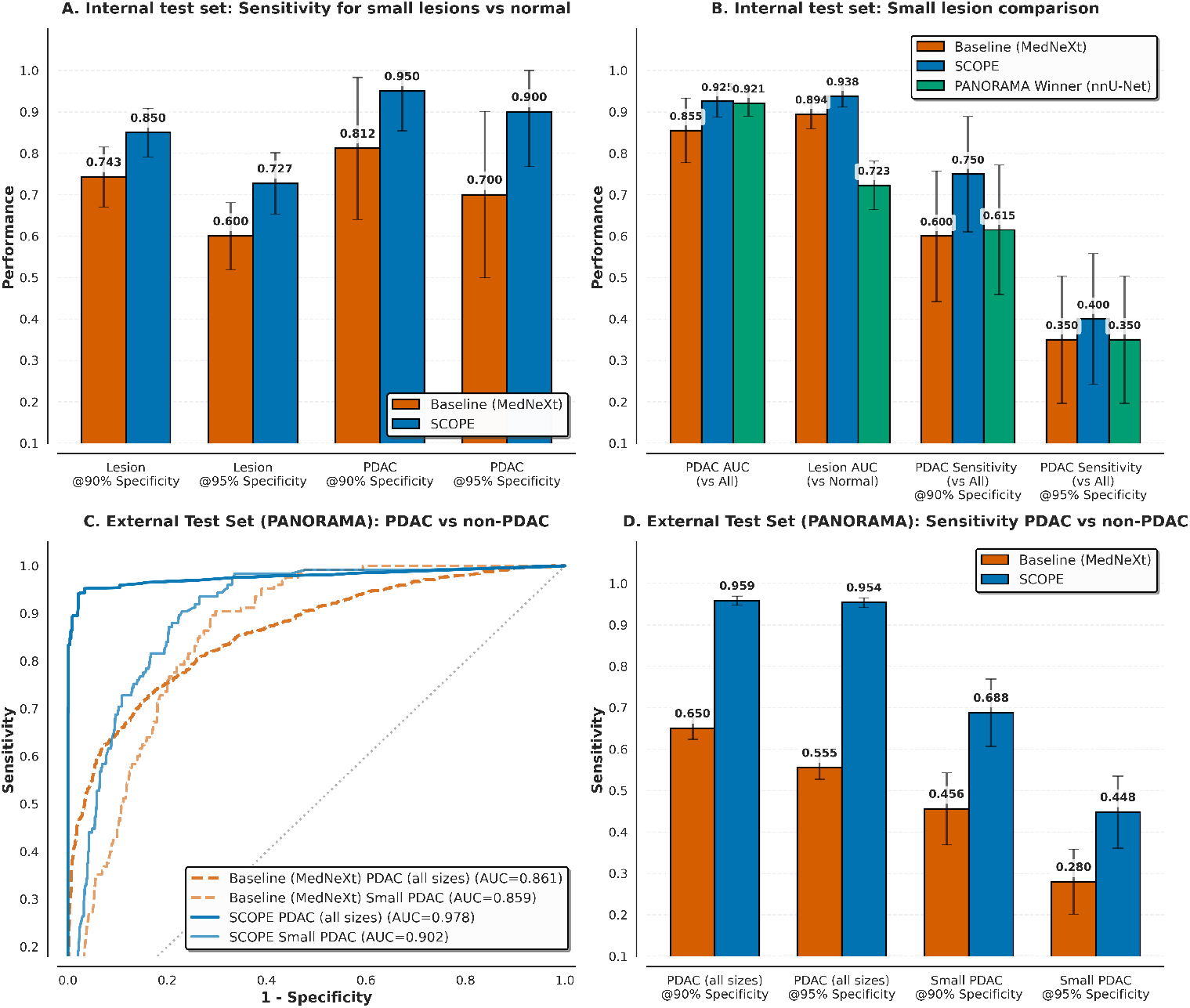
Performance in internal and external test sets. **A)** SCOPE sensitivity at high specificity operating points in internal test set. **B)** Model performance in the internal test set, including the PANORAMA winner model (PanDx) for PDAC vs non-PDAC detection. **C)** Receiver operating characteristic curves in the external test set. **D)** Model sensitivity at high specificity in the external test set, considering PDAC of all sizes and small size. Error bars are 95% confidence intervals of the mean.

To benchmark against a state-of-the-art PDAC model, we evaluated the pre-trained PANORAMA winner (PanDx) (26) (nnU-Net) on the internal test set (Figure 3B). For small PDAC in the vs-all setting, SCOPE achieved an AUC of 0.925 (95% CI: 0.888, 0.963), statistically non-inferior to PanDx (0.921; non-inferiority *P* < .001). Sensitivity at 90% specificity (0.750 vs 0.615) and 95% specificity (0.400 vs 0.350) further confirmed non-inferiority (*P* = .003, *P* = .015). In contrast, in the PDAC-vs-normal task, SCOPE achieved significantly higher performance than PanDx (no retraining) with sensitivities of 0.950 vs 0.654 (*P* < .001) and 0.900 vs 0.624 (*P* < .001) at 90% and 95% specificity, respectively. For detecting any small lesion versus normal, SCOPE again strongly outperformed PANORAMA (AUC = 0.938 [95% CI: 0.912, 0.964] vs 0.723; *P* < .001), underscoring that specialized PDAC vs non-PDAC models may generalize poorly to multi-lesion tasks and are highly dependent on how the non-PDAC class is defined. Table 2 summarizes these findings (ablation study in Table S3).

### 3.2 External multi-institutional test set

On the external 2,238-case dataset from the PANORAMA challenge (training cohort), SCOPE showed large improvements over the baseline segmentation model (MedNeXt) without retraining. For PDAC of all sizes versus non-PDAC (Figure 3C), SCOPE achieved an AUC of 0.978 superior to 0.861 for the baseline, and for small PDAC, SCOPE again outperformed the baseline (0.902 vs 0.859) with *P* < .001 in both cases. At the high specificity operating points (Figure 3D), for all PDAC cases, SCOPE sensitivity at 90% specificity was 0.959 (95% CI: 0.947, 0.970) vs 0.650 for the baseline and at 95% specificity was 0.954 (95% CI: 0.942, 0.966) vs 0.555, *P* < .001 in both. For small PDAC cases, SCOPE sensitivity at 90% specificity improved from 0.456 to 0.688 (95% CI: 0.607, 0.769; *P* = .028), and at 95% specificity improved from 0.280 to 0.448 (95% CI: 0.361, 0.535; *P* = .039).

### 3.3 Small-lesion reader study

On the small-lesion reader study (40 PDACs, 40 pancreatic cysts, 20 PanNETs, and 100 normal controls), three abdominal radiologists demonstrated high lesion-vs-normal sensitivity with moderate specificity (Figure 4C). Most reader errors reflected subtype misclassification (e.g., PDAC confused with cyst) or false positives rather than missed lesions, with normal cases frequently classified as cyst or PanNET. Per-class performance showed substantial inter-reader variability, particularly for PanNET, and more homogeneous performance for PDAC and cysts (Figure 4B–C).

**Figure 4:**
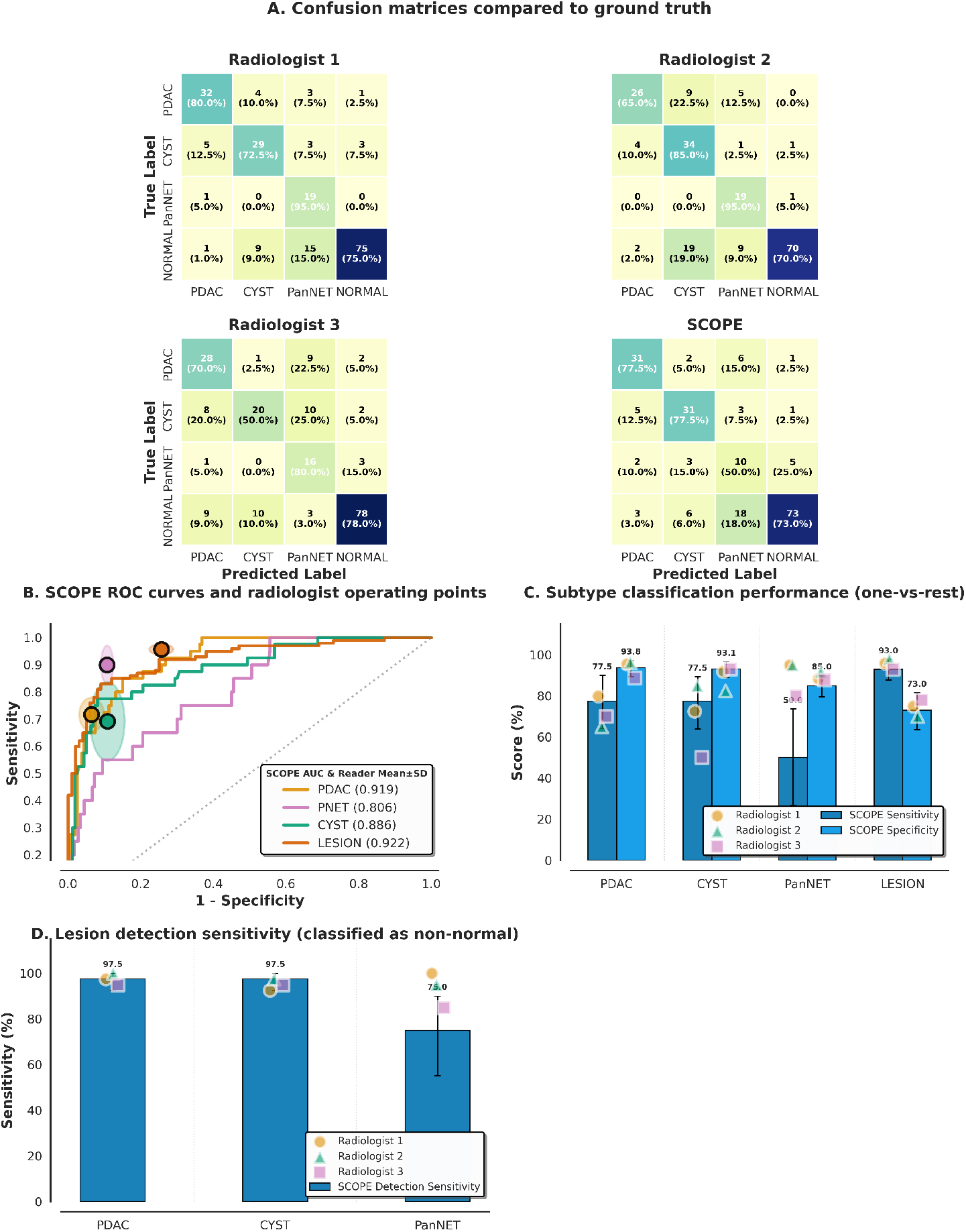
Small lesion reader study. **A)** Confusion matrices for radiologists and model compared to ground truth. **B)** Receiver operating curves and radiologist operating points. **C)** Classification performance vs all other cases. **D)** Lesion detection sensitivity per class, defined as the proportion of cases of a given lesion type that are detected as non-normal.

**Figure 5:**
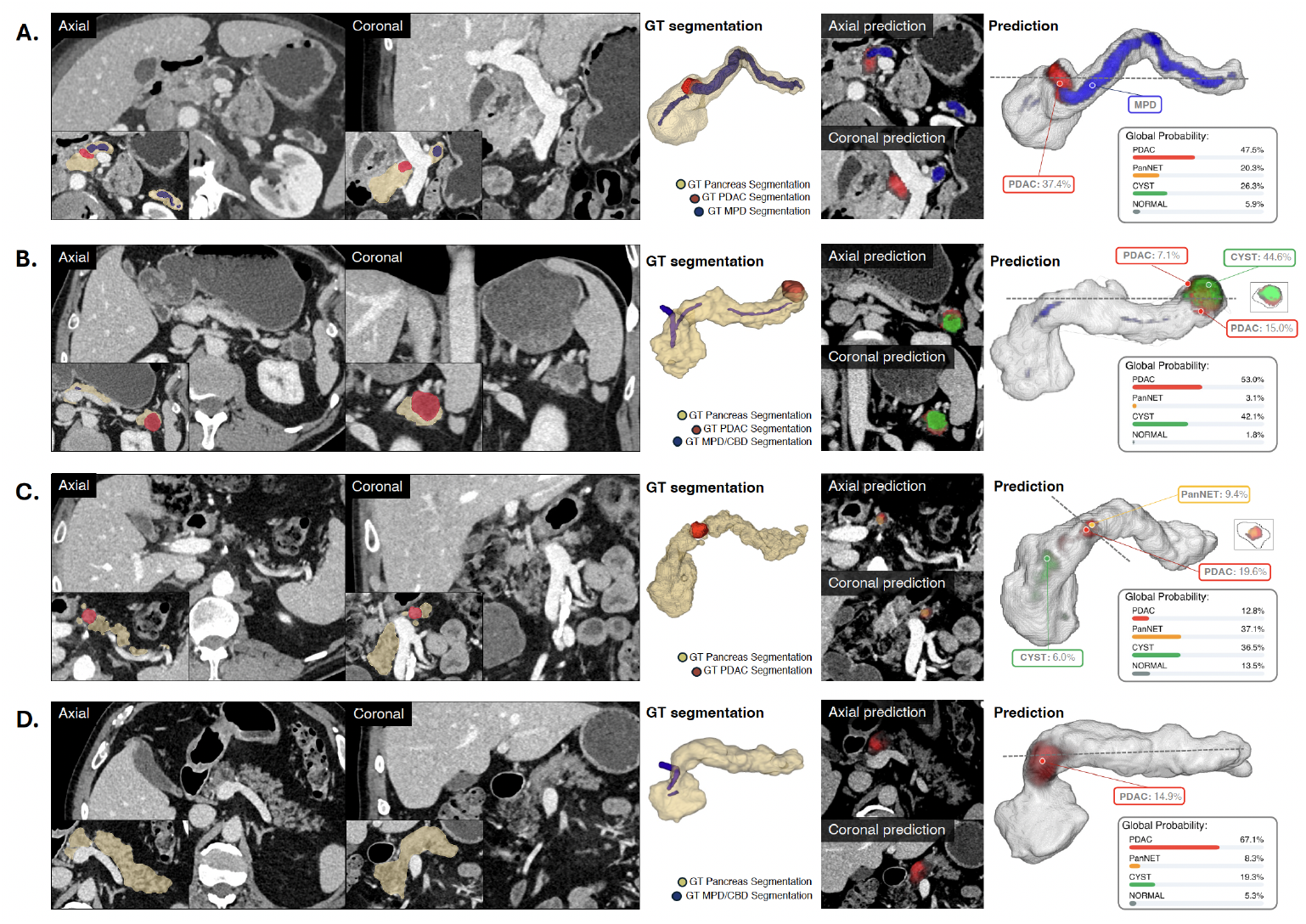
Ground truth compared to SCOPE predictions, including global probability computed by the metamodel. (A) Biopsy-proven PDAC with main pancreatic duct dilation and abrupt duct cutoff; all readers and SCOPE correctly classified PDAC, with SCOPE highlighting the lesion region and ductal abnormality. (B) Biopsy-proven PDAC with mixed morphology; two readers misclassified the case as PanNET and cyst, whereas SCOPE delineated cystic (green) and solid (red) components with ductal dilation and correctly classified the case as PDAC. (C) Biopsy-proven PDAC in the pancreatic body; SCOPE localized the lesion with local prediction favoring PDAC but global misclassification as PanNET. Two readers also classified the case as PanNET, and the third as cyst. (D) No known pancreatic pathology; SCOPE produced a false-positive PDAC prediction in the setting of parenchymal attenuation heterogeneity, while all readers classified the case as normal. *MPD*: Main Pancreatic Duct; *GT*: Ground truth; *CBD*: Common Bile Duct. Colors in “Global Probability” match those presented in voxel-level predictions.

**Figure 6:**
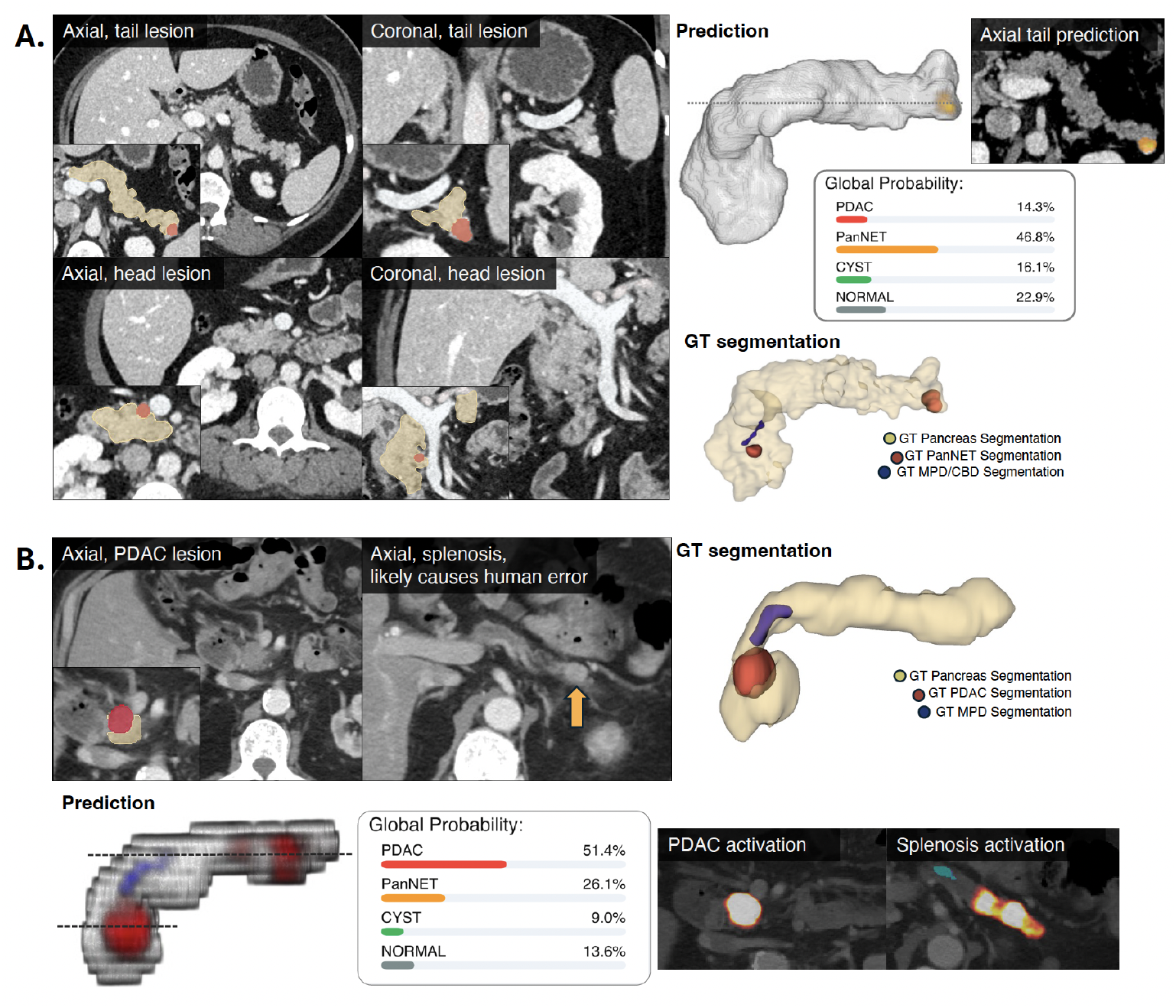
SCOPE predictions compared to ground truth, including global probability computed by the metamodel. (A) Two hypervascular, histopathology-confirmed PanNETs (tail 8 mm; head 5 mm); all readers and SCOPE classified the case as PanNET, although SCOPE detected only the larger tail lesion. (B) Biopsy-proven PDAC on an outside lower-resolution CT (5-mm slice thickness; axial views shown to readers); a splenosis focus (orange arrow) was present in the setting of prior splenectomy. All readers classified the case as PanNET, whereas SCOPE classified PDAC and highlighted both the PDAC and splenosis as candidate lesions in its local predictions. *MPD*: Main Pancreatic Duct; *GT*: Ground truth; *CBD*: Common Bile Duct. Colors in “Global Probability” match those presented in voxel-level predictions.

SCOPE, trained exclusively on the internal training set and using a metamodel for mutually exclusive classification, achieved a lesion-vs-normal AUC of 0.922 (95% CI: 0.882, 0.955), and vs-all AUCs of 0.919 (95% CI: 0.874, 0.953) for PDAC, 0.886 (95% CI: 0.815, 0.942) for cysts, and 0.806 (95% CI: 0.698, 0.892) for PanNETs (Figure 4B). At its operating point, SCOPE had competitive performance for all classes except PanNET. The model confusion matrix shows that SCOPE correctly identified 31/40 PDACs (77.5%), 31/40 cysts (77.5%), and 10/20 PanNETs (50%), with high specificities for each class (0.85–0.93). Statistical tests are presented in Table S4.

Across cases with at least one incorrect reader interpretation, SCOPE correctly classified 68 cases (34.0% [68/200] of all cases), including 29 cases (14.5%) with two or more incorrect readers and 9 cases (4.5%) misclassified by all three readers (Figure S1); additionally, among lesion cases, SCOPE correctly identified 8.0% (8/100) of lesions missed by at least one reader. When the task was defined as non-normal detection (Figure 4D), irrespective of lesion subtype, SCOPE detected 97.5% of PDAC and cyst cases and 75% of PanNET cases, with 25% (5/20) of PanNET cases incorrectly predicted as normal.

### 3.4 Comparison to past studies

Table 3, adapted from Lopez–Ramirez et al. (5), summarizes relevant prior AI studies. Most published work includes relatively few small lesions and predominantly evaluates PDAC detection versus normal controls. In contrast, the present study evaluates one of the largest cohorts of small pancreatic lesions to date, spanning multiple lesion types and including a dedicated reader study. Although task definitions, negative-class composition, and operating points vary widely across studies, SCOPE demonstrated high sensitivity both for the common PDAC-versus-normal task and for class-wise detection of small lesions across PDAC, cysts, and PanNETs.

**Table 3:**
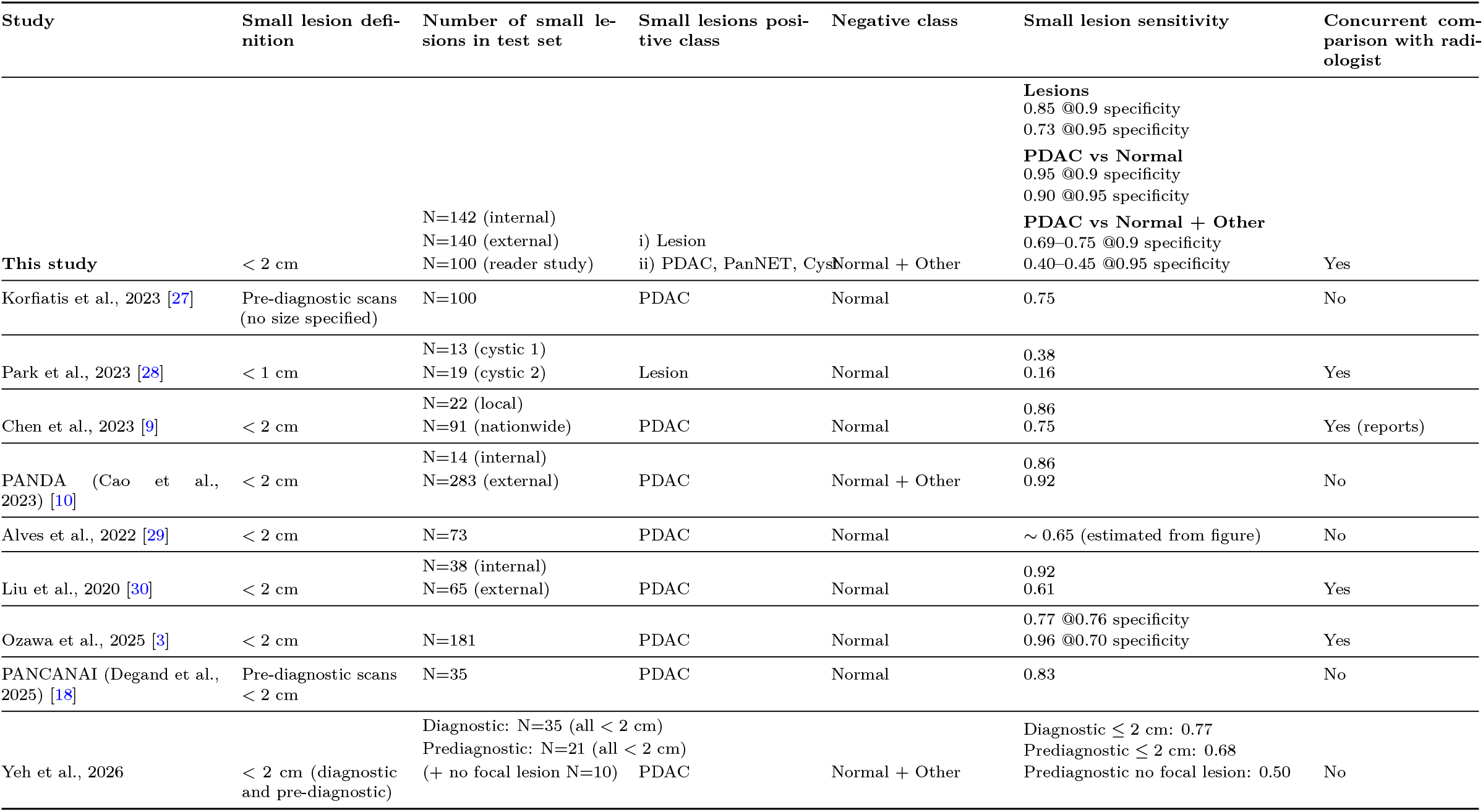
Comparison of this study to prior studies reporting stratified results for lesions ≤ 2 cm.

## 4 Discussion

In this study, we developed SCOPE, a deep learning model that integrates voxel-level features with global pancreatic context to improve the detection of small pancreatic lesions in CT. Across an internal cohort, a multi-institutional external PDAC dataset, and a curated small-lesion reader study, SCOPE consistently outperformed a strong segmentation baseline and matched or exceeded PDAC-specific state-of-the-art performance while extending robust detection to cysts and PanNETs. SCOPE improved internal small-lesion AUC from 0.956 to 0.974, increased small-lesion sensitivity at 95% specificity from 0.600 to 0.727, and raised PDAC AUC in the external PANORAMA cohort from 0.861 to 0.978 without retraining. In the small-lesion reader study, SCOPE achieved a lesion-vs-normal AUC of 0.922 and performed within the range of experienced abdominal radiologists. These findings support the central hypothesis that combining local and global information improves small lesion detection and classification.

Small pancreatic lesions remain difficult to detect because early PDAC, and some PanNETs, often lack a discrete mass and instead present through subtle, spatially distributed abnormalities such as focal duct cutoff, asymmetric parenchymal texture, or mild gland atrophy; small cystic lesions may also be inconspicuous, particularly when lacking overt ductal dilation or wall thickening. Prior AI approaches—trained largely as mass detectors on simplified PDAC-vs-normal or narrow PDAC-vs-non-PDAC tasks—typically underperform for sub-2 cm lesions (Table 3). In contrast, SCOPE incorporates explicit segmentations of the pancreatic parenchyma, ducts, and lesions alongside whole-gland context. Its strong performance under the more realistic versus-all evaluation, where lesions must be distinguished not only from normal pancreas but also from other explicit lesion types, together with the marked performance drop observed for PDAC-specialized models when the negative class is broadened, highlight two requirements for clinical validation: global-context modeling and careful definition of the negative class.

Architecturally, SCOPE improves upon prior segmentation–classification approaches by pairing a transformer-based encoder with a training strategy that avoids degrading anatomical segmentations. This balance between segmentation fidelity and long-context is consistent with emerging encoder architectures for medical imaging (31). In addition, a lightweight metamodel allowed SCOPE to adapt its outputs to different diagnostic tasks—including PDAC-versus-non-PDAC detection, lesion triage, or multi-class classification—without retraining the backbone, offering practical flexibility for deployment across diverse institutional workflows.

The three complementary test sets clarify SCOPE’s clinical potential. In the internal dataset, contextual modeling improved detection across lesion types in a controlled, fully annotated environment. In the external dataset, these gains generalized across institutions and scanners. In the small-lesion reader study, SCOPE achieved expert-level sensitivity for most tasks while maintaining high specificity. Here, SCOPE correctly classified 15.0% of cases that two or more readers misclassified, supporting potential use as a decision-support tool. As almost all sub-2 cm PanNETs and cysts are curable, and most node-negative sub-2 cm PDACs are curable, detection of these lesions has the potential to identify curable neoplasms before they progress to incurable metastatic cancers (32,33). If applied in the background to the many CT scans obtained for another indication, this approach would not entail additional radiation. Furthermore, focusing on high-risk patients, such as those with new-onset diabetes after age 50, a family history of pancreatic cancer, or unexplained weight loss and back pain (34), SCOPE has the potential to identify a reasonable number of asymptomatic, potentially treatable lesions with a low false-positive rate.

This study has limitations. PanNET sensitivity remained lower than for PDAC and cysts, reflecting limited diversity in the training set and the difficulty of detecting small hypervascular tumors on venous-phase CT. In addition, only venous-phase scans were used for model development; arterial CT or MRI may provide additional discriminatory information for certain lesion subtypes. Furthermore, although SCOPE provides both voxel-level and global predictions, the optimal aggregation, calibration, and presentation of these signals for radiologist interpretation remain open challenges. Finally, this was a retrospective study; prospective evaluation in well-defined high-risk or screening populations (35) will be required for workflow improvements and clinical utility.

In conclusion, SCOPE has the potential to advance early pancreatic lesion detection by combining local segmentation with global pancreatic context, enabling more reliable identification of small PDACs, cysts, and PanNETs across clinically diverse settings. Its consistent performance across internal, external, and expert-reader cohorts, together with task flexibility, supports its potential role as a decision-support tool to assist radiologists in earlier and more accurate detection of pancreatic neoplasms. Prospective validation, evaluation in pre-diagnostic imaging, and integration of clinical and radiomic features represent important next steps toward clinical translation.

## Data Availability

All data produced in the present study are available upon reasonable request to the authors

