## Supplemental Information for "SCOPE: AI-Assisted Early Detection of Potentially Curable Pancreatic Neoplasms on CT from Local and Global Information"

#### Appendix S1. CT acquisition and data preparation

##### Internal training and test set

The cohort comprised 4,065 dual-phase contrast-enhanced abdominal CT examinations acquired in a single institution in the period 2003-2024. Examinations were predominantly performed on Siemens Healthineers platforms (3,903/3,965; 98.4%), most commonly SOMATOM Definition Flash (1,648; 41.6%), SOMATOM Force (737; 18.6%), Sensation 64 (407; 10.3%), SOMATOM Drive (345; 8.7%), and SOMATOM Definition (248; 6.3%). A minority of studies were acquired on scanners from GE Medical Systems (0.7%), Philips (0.6%), and Toshiba (0.3%). Tube potential was typically 120 kVp (median 120, IQR: 100–120, range: 80–140), with 68.5% of examinations acquired at 120 kVp. Lower tube potentials (90–100 kVp) accounted for approximately 24% of studies, consistent with dose-optimized abdominal imaging protocols. Reconstruction was performed at thin section thicknesses, with a median slice thickness of 0.5 mm (IQR, 0.5–0.5; range, 0.5–5.0 mm), and 90.5% of volumes reconstructed at 0.5 mm.

In-plane spatial resolution was high, with a median pixel spacing of 0.73 mm (IQR, 0.67–0.79 mm; range, 0.33–1.00 mm), yielding near-isotropic voxels in most cases. Detector collimation metadata were available for 3,149/3,965 (79.4%) examinations. Among these, single collimation was most frequently 0.6 mm (96.0%), and total collimation was most commonly 38.4 mm (65.8%), with a broader range spanning 10–80 mm, reflecting scanner- and protocol-dependent acquisition configurations. Image reconstruction kernels were predominantly soft-tissue kernels optimized for abdominal imaging. The most frequently used kernels included I26f (34.2%), B20f (24.5%), Br36d (18.3%), B30f (8.5%), and Br36f (7.0%), with remaining kernels collectively accounting for less than 8% of reconstructions. Bowtie filtration was documented as wedge-type in 41.8% of scans and flat filtration in 33.4%, while filtration type was unspecified in 23.2% of examinations.

##### Diagnostic reference standards and spatial ground truth

An earlier version of this dataset has been described in a previous study [Anonymized Reference]. Diagnostic labels for pancreatic disease were established using clinical reference standards available at the time of imaging. PDAC and PanNET diagnoses were confirmed by histopathology, obtained from surgical resection specimens or image-guided biopsy, and cross-referenced with pathology reports. Pancreatic cysts were defined based on a combination of radiologic imaging features, clinical records, and, where available, pathologic or cytologic confirmation. Normal control cases were derived primarily from renal donor populations and patients without known pancreatic disease, with absence of pancreatic pathology confirmed through clinical history and longitudinal follow-up (2 years), when available. All diagnostic labels were assigned retrospectively and independently of the spatial annotation process.

Spatial ground truth was established through manual three-dimensional delineation of the pancreas and pancreatic lesions across the full volumetric extent of each CT examination. Annotations were performed by five trained annotators using commercial medical image annotation software, with explicit attention to

Classified as Microsoft Confidential

lesion localization, spatial extent, and boundary definition in axial, coronal, and sagittal planes. For subjects with dual-phase (arterial and venous) imaging, pancreatic structures and lesions were annotated separately in each phase to preserve phase-specific spatial characteristics.

To ensure spatial accuracy and consistency, all annotations underwent independent secondary review by one of three board-certified abdominal radiologists who did not participate in the initial annotation or the subsequent reader study. A structured quality-assurance pipeline was applied to identify potential spatial inconsistencies, including incomplete organ coverage, discontinuous boundaries, or implausible lesion geometry. Automated checks using in-house software quantified region-of-interest metrics and flagged outliers for further review. Cases requiring correction—such as small lesions initially missed or minor spatial misregistrations—were revised following targeted radiologist re-evaluation.

### Appendix S2. SCOPE Model Development

#### Network architecture

SCOPE is a multi-head 3D model comprising (i) a volumetric encoder-decoder for multi-class segmentation and (ii) a parallel case-level classification head that fuses encoder-derived contextual features with segmentation-derived structural signals (Figure 1B). All networks operated on pancreas-localized, cropped volumes (PanSegNet) and produced predictions over overlapping 3D regions of interest (ROIs), which were aggregated to obtain case-level outputs.

Each forward pass of SCOPE consumed a single-channel 3D patch, $x \in \mathbb{R}^{Bx96\times96\times96}$, and produced both segmentation logits, $seg_{out} \in \mathbb{R}^{7\times96\times96\times96}$ (background, pancreas, PDAC, PanNET, cyst, main pancreatic duct, common bile duct) and case-level classification logits $cls_{out} \in \mathbb{R}^{B\times4}$ (normal, PDAC, cyst, PanNET).

##### Encoder and segmentation decoder

For the encoder and the segmentation head we used the SwinUNETR backbone (MONAI) instantiated with img_size = (96,96,96), in_channels = 1, out_channels = 7, feature_size = 48, depths = (2,2,2,2), num_heads = (3,6,12,24), drop_rate = attn_drop_rate = dropout_path_rate = 0, normalize = True, and use_checkpoint = True (spatial_dims = 3). The implementation used a patch size of (2,2,2) and a window size of (7,7,7). Results using this encoder and segmentation decoder are presented in the main manuscript.

##### Classification head

For the classification head, we used the 192 channel-encoder feature maps (encoder_3), which provided a favorable trade-off between spatial detail and semantic abstraction, while keeping the fused classifier input compact. This map was passed to a coordinate attention mechanism (1). Let $x \in \mathbb{R}^{B\times192\times D'\times H'\times W'}$ denote this input to the classification head. Axis-wise average pooling was first applied independently along each spatial dimension to encode long-range directional context:

Classified as Microsoft Confidential

$$z_d = \text{AvgPool}(x) \in R^{B \times C \times D' \times 1 \times 1}$$

$$z_h = \text{AvgPool}(x) \in R^{B \times C \times 1 \times H' \times 1}$$

$$z_w = \text{AvgPool}(x) \in R^{B \times C \times 1 \times 1 \times W'}$$

Each pooled tensor was independently projected using a 1×1×1 convolution, followed by batch normalization and ReLU activation, reducing the channel dimensionality 8. Each branch was then expanded back to C = 192 channels via a second 1×1×1 convolution and passed through a sigmoid nonlinearity to produce axis-specific attention maps $a_d$, $a_h$, and $a_w$. The refined encoder feature map was obtained by multiplicative gating:

$$x' = x \odot a_d \odot a_h \odot a_w$$

This operation enables the model to encode long-range dependencies along each spatial axis while preserving precise localization cues relevant to subtle pancreatic morphologic changes.

The classifier fused global contextual information from the encoder with local information from segmentation outputs. Global average pooling (GAP) was applied to both the refined encoder feature map and the segmentation logits:

$$f_{enc} = \text{GAP}(x') \in \mathbb{R}^{B \times 192}$$

$$f_{seg} = \text{GAP}(\text{seg}_{\text{out}}) \in \mathbb{R}^{B \times 7}$$

The pooled features were concatenated to form a fused representation $\in \mathbb{R}^{B \times 199}$ and passed to a two-layer fully connected network:

$$\text{Linear}(199 \to 128) \to \text{BatchNorm} \to \text{ReLU} \to \text{Dropout}(p = 0.3) \to \text{Linear}(128 \to 4),$$

producing logits for normal, PDAC, cyst, and PanNET for a given patch or ROI.

Finally, the classification logits for multiple patches in the pancreas are aggregated using max pooling, independently for each diagnostic class. After applying a sigmoid activation, this produces a final global classification probability for each case for normal, PDAC, cyst, and PanNET, matching case-level supervision.

**Backbone variant, SCOPE-MedNeXt**

As an additional evaluated variant (SCOPE-MedNeXt**)**, we replaced the SwinUNETR encoder–decoder with a MedNeXt-M segmentation backbone while keeping the same overall multi-head design and training schedule. The MedNeXt backbone was instantiated with spatial_dims=3, in_channels=1, out_channels=7, and init_filters=48, using blocks_down=(2,2,2,2), blocks_bottleneck=2, and blocks_up=(2,2,2,2), with group normalization and residual connections. The attached classification head used the MedNeXt bottleneck feature map with C=768 channels as classification head input, i.e., $x \in \mathbb{R}^{B \times 786 \times D' \times H' \times W'}$, and otherwise matched the coordinate-attention + pooling + MLP formulation described above. Model performance was competitive with SwinUNETR, both we noted lower performance in the internal test sets and reader study cohort.

Classified as Microsoft Confidential

**Metamodel**

The SCOPE training architecture is designed multi-label predictions across a variety of classes. However, depending on the class definitions and target tasks, the aggregation of local and global features might be considered different for a final decision.

Thus, in some tasks, case-level predictions were done using a lightweight metamodel trained with FLAML AutoML (2) on structured outputs derived from SCOPE inference. For each case, the metamodel used a feature vector of: (i) the case-level classification per-class probabilities from the classification head, and (ii) statistics from the post-processed segmentation probability maps, including the maximum and non-zero mean probability values for PDAC, pancreatic cyst, PanNET, pancreatic duct, and common bile duct. These features capture complementary global diagnostic confidence and localized structural evidence without requiring retraining of the neural network for other tasks or class definitions. For the results of the study, two metamodels were trained exclusively on the internal training set, *PDAC vs non-PDAC* (used for predictions on the PANORAMA external test set) and mutually exclusive PDAC, cyst, PanNET and normal (used for the reader study predictions).

### Data pipeline and model training

**Preprocessing**

All CT volumes were localized and cropped to the pancreas region using PanSegNet and reoriented to RAS. Each patch was resampled to spacing of 1.5×1.5×1.5 mm (bilinear for images; nearest neighbor for labels), and then clipped to the range [-100, 200] and linearly scaled to 0, 1. From this cropped volume, patches of fixed-size of 96×96×96 voxels where sampled and padded as appropriate.

During training, weighted sampling was performed at ration of 9:1 non-zero to background patches, with 2 sample per iteration. On-the-fly 3D augmentations were applied during training: 1) Random 90° rotations with probability 0.8, max_k = 3, ii) random flip with probability 0.4, iii) intensity shift with probability 0.5 and offsets up to ±0.10, iv) Gaussian smoothing with probability 0.2, σ sampled in [0.5, 1.15] for each axis, v) Gaussian noise: probability 0.2, σ=0.01.

**Training schedule and optimization**

The training followed the three-stage procedure described in the main manuscript:

1) Segmentation warm-up: encoder and decoder trained from scratch for 1,000 epochs using dice cross entropy loss (DiceCE) for multi-class segmentation and a learning rate of $1\times10^{-4}$.

Classified as Microsoft Confidential

2) Classification warm-up: with encoder and decoder frozen, the classification head was trained for 75 epochs using multi-label binary cross-entropy (BCE) loss, applied independently to each diagnostic class. For each case, patch-level logits were aggregated via max pooling across ROIs to obtain case-level logits, which were transformed using independent sigmoid functions. As described in the model architecture, the per-patch sigmoid probability per class was aggregated by taking the maximum across multiple patches, producing probabilities at the case level. A learning rate of $1\times10^{-3}$ was used.

3) Joint fine-tuning: in the final stage, all network parameters were unfrozen, and the model was jointly optimized for segmentation and classification. The total loss was defined as a weighted sum of the voxel-level segmentation loss and the case-level classification loss:

$$\mathcal{L} = \alpha\,\mathcal{L}_{seg} + \beta\,\mathcal{L}_{cls}$$

where $\mathcal{L}_{seg}$ denotes the voxel-wise cross entropy loss, and $\mathcal{L}_{cls}$ denotes the multi-label binary cross-entropy loss. We used weights $\alpha = 1.0$ and $\beta = 0.1$ to prioritize segmentation performance. We found two important parameters maximize model performance: 1) we use cross entropy loss instead of DiceCE loss to facilitate lesion detection, as done in (3), 2) the gradients from the classification loss can degrade the segmentation performance during optimization. To avoid this latter problem, we prevented the classification gradients from backpropagating into the segmentation decoder, only updating the encoder and classification head with the classification loss (usually referred as gradient detaching). A learning rate of $1\times10^{-4}$ was used for 400 epochs.

All optimizations used AdamW with weight decay of $1\times10^{-5}$ and a linear warmup (5 epochs) plus cosine annealing learning-rate schedule. Hyperparameter tuning was performed by grid search of most sensitive parameters, including learning rate of stage 3 tuning [$1\times10^{-4}$, $5\times10^{-4}$ , $1\times10^{-3}$], $f_{enc}$ dimensionality on the of the classification attention head and discrete $\alpha, \beta$ configurations; performance was selected based on 10% validation test set sampled randomly from the training set, and the re-trained on the complete internal training set. Distributed data parallel training and automated mixed precision (AMP) were used for efficiency and scaling. Using 4 A100 GPUs, SCOPE took 188 hours for stage 1 training, 5 hours for stage 2, and 76 hours for stage 3.

The metamodel, trained on the internal training set, was used for predictions in the external test set and the reader study. FLAML used a fixed time budget of 180 seconds, classification task option, accuracy optimization metric, with candidate estimators limited to gradient-boosted decision trees (LightGBM), random forest, and L1-regularized logistic regression.

**2.2.5 Postprocessing**

Classified as Microsoft Confidential

Post-processing was performed using a static thresholding strategy with class-specific probability thresholds of 0.5 for pancreas, 0.3 for pancreatic duct and common bile duct, and very permissive 0.01 for PDAC, pancreatic cyst, and PanNET, followed by removal of connected components smaller than 100 voxels. Candidate lesion regions were required to be adjacent to the pancreas and segmented ducts, enforced via morphological dilation–based adjacency, to ensure anatomical plausibility. This post-processing pipeline reduced small, isolated false positives while preserving sensitivity to small focal pancreatic lesions prior to feature extraction for the metamodel.

**Benchmark models**

To contextualize SCOPE performance, we trained and evaluated several segmentation-only benchmark models using identical data preprocessing, pancreas localization, ROI extraction, and evaluation protocols. The benchmark architectures included MedNeXt, SwinUNETR, and nnU-Net, all trained from scratch on the same segmentation targets (pancreas, PDAC, pancreatic cyst, PanNET, pancreatic duct, and common bile duct). An additional specialized baseline, the pretrained PANORAMA winner (PanDx, using nnU-Net), was evaluated without retraining and used only for PDAC versus non-PDAC detection comparisons.

For MedNeXt, SwinUNETR, and nnU-Net, models were trained for 1,000 epochs using a DiceCE objective on voxel-level segmentation outputs, with a learning rate of $1\times10^{-4}$, linear warm-up, and cosine annealing learning-rate schedule, matching the SCOPE training pipeline. For MedNeXt, we used the variation MedNeXt-M, as the larger MedNeXt-L variant was excluded due to substantially higher computational cost without measurable performance gains in preliminary experiments.

Model selection among the segmentation-only baselines was based on small-lesion AUC in the internal test set, reflecting the primary clinical objective of early lesion detection. MedNeXt-M achieved the highest small-lesion AUC and was therefore chosen as the primary segmentation benchmark for reporting. Notably, MedNeXt-M also demonstrated superior performance for larger lesions across all classes compared with SwinUNETR and nnU-Net, while the PANORAMA (PanDx) model served as a strong task-specific benchmark for PDAC detection.

### Appendix S3. Supplemental Results

**Choosing SCOPE segmentation backbone**

We chose SCOPE segmentation backbone based on performance in the internal test set. Table S1 presents performance metric according to the SCOPE segmentation backbone. SwinUNETR was the best performing backbone, and it is used in all results in the main manuscript.

*Table S1: SCOPE performance* ***on small lesions*** *in the internal test. The most stringent condition (vs All) is used to determine the best backbone. SwinUNETR was the best performing backbone as is used in all results in the main manuscript.*

| SCOPE Backbone | Metric | Value |
|---|---|---|
| **SwinUNETR** | **Lesion AUC** | **0.938 [0.912, 0.964]** |

Classified as Microsoft Confidential

| | | |
|---|---|---|
| MedNeXt | Lesion AUC | 0.919 [0.899, 0.941] |
| **SwinUNETR** | **PDAC AUC (vs All)** | **0.925 [0.888, 0.963]** |
| MedNeXt | PDAC AUC (vs All) | 0.882 [0.858, 0.924] |
| **SwinUNETR** | **PanNET AUC (vs All)** | **0.907 [0.850, 0.963]** |
| MedNeXt | PanNET AUC (vs All) | 0.851 [0.826, 0.890] |
| **SwinUNETR** | **Cyst AUC (vs All)** | **0.903 [0.869, 0.937]** |
| MedNeXt | Cyst AUC (vs All) | 0.887 [0.838, 0.922] |

**Dice Scores**

*Table S2: Dice Similarity Coefficient for SCOPE predictions on the internal test set. No overlap was penalized as DSC=0.*

| ***Class*** | ***Cohort*** | ***Mean DSC*** | ***95% CI*** | ***N*** |
|---|---|---|---|---|
| PDAC | All lesions | 0.574 | [0.535, 0.612] | 207 |
| PNET | | 0.662 | [0.597, 0.727] | 100 |
| PAN-CYST | | 0.555 | [0.502, 0.609] | 149 |
| Any lesion | | 0.611 | [0.583, 0.639] | 429 |
| PDAC | Small lesions | 0.285 | [0.156, 0.413] | 20 |
| PNET | | 0.485 | [0.367, 0.604] | 41 |
| PAN-CYST | | 0.488 | [0.416, 0.560] | 81 |
| Any lesion | | 0.493 | [0.437, 0.548] | 140 |

**Ablation study**

Based on the SCOPE architecture, we considered two model variations: (1) Segmentation backbone only without a classification head, (2) No coordinate attention in classification head, (3) No gradient flow blocking into segmentation decoder. We used small lesion AUC as the metric of interest, as presented in Table S3, as it found to be a consistent proxy of small lesion performance across all classes of interest and consistent with vs all and vs normal per-class performance in most cases.

*Table S3: SCOPE ablation study, in all cases all other model and training configurations remain unchanged. Metrics are evaluated in the internal test set.*

| SCOPE Variation | Metric | Value |
|---|---|---|
| **Final model** | **Small lesion AUC** | **0.938 [0.912, 0.964]** |
| No classification head (SwinUNETR backbone) | Small lesion AUC | 0.863 [0.818, 0.901] |
| No coordinate attention in classification head | Small lesion AUC | 0.898 [0.868, 0.931] |

Classified as Microsoft Confidential

| | | |
|---|---|---|
| No gradient flow blocking into segmentation decoder | Small lesion AUC | 0.913[0.885, 0.944] |

**Statistical tests**

The following tables present results of superiority and non-inferiority statistical test for comparisons reported in the manuscript.

*Table S4: Statistical tests and their results for Table 2. Values in bold corresponds to those test that statistically significant with P < 0.05. All results are for small lesions in the internal test set.*

| Metric | Target | Comparison | Setting | Test Type | Superiority p-value | Non-inferiority p-value |
|---|---|---|---|---|---|---|
| AUC | Lesion | SCOPE vs Baseline | vs Normal | DeLong | **p=0.005** | **p<0.001** |
| AUC | Lesion | SCOPE vs PANORAMA | vs Normal | DeLong | **p<0.001** | **p<0.001** |
| AUC | PDAC | SCOPE vs Baseline | vs Normal | DeLong | **p=0.012** | **p<0.001** |
| AUC | PDAC | SCOPE vs Baseline | vs All | DeLong | **p=0.004** | **p<0.001** |
| AUC | PDAC | SCOPE vs PANORAMA | vs Normal | DeLong | p=0.075 | **p<0.001** |
| AUC | PDAC | SCOPE vs PANORAMA | vs All | DeLong | p=0.382 | **p<0.001** |
| AUC | PanNET | SCOPE vs Baseline | vs Normal | DeLong | **p=0.004** | **p<0.001** |
| AUC | PanNET | SCOPE vs Baseline | vs All | DeLong | **p=0.007** | **p<0.001** |
| AUC | Cyst | SCOPE vs Baseline | vs Normal | DeLong | p=0.238 | **p=0.002** |
| AUC | Cyst | SCOPE vs Baseline | vs All | DeLong | p=0.447 | **p<0.001** |
| Sensitivity @ 90% Specificity | Lesion | SCOPE vs Baseline | vs Normal | McNemar | **p=0.047** | **p<0.001** |
| Sensitivity @ 90% Specificity | Lesion | SCOPE vs PANORAMA | vs Normal | McNemar | **p<0.001** | **p<0.001** |
| Sensitivity @ 90% Specificity | PDAC | SCOPE vs Baseline | vs Normal | McNemar | **p<0.001** | **p=0.010** |
| Sensitivity @ 90% Specificity | PDAC | SCOPE vs Baseline | vs All | McNemar | **p<0.001** | **p=0.037** |
| Sensitivity @ 90% Specificity | PDAC | SCOPE vs PANORAMA | vs Normal | McNemar | **p<0.001** | **p=0.002** |
| Sensitivity @ 90% Specificity | PDAC | SCOPE vs PANORAMA | vs All | McNemar | **p=0.136** | **p=0.003** |
| Sensitivity @ 90% Specificity | PanNET | SCOPE vs Baseline | vs Normal | McNemar | **p=0.024** | **p<0.001** |
| Sensitivity @ 90% Specificity | PanNET | SCOPE vs Baseline | vs All | McNemar | **p=0.031** | **p=0.001** |
| Sensitivity @ 90% Specificity | Cyst | SCOPE vs Baseline | vs Normal | McNemar | p=0.721 | p=0.376 |
| Sensitivity @ 90% Specificity | Cyst | SCOPE vs Baseline | vs All | McNemar | p=0.729 | **p=0.025** |
| Sensitivity @ 95% Specificity | Lesion | SCOPE vs Baseline | vs Normal | McNemar | **p=0.006** | **p<0.001** |
| Sensitivity @ 95% Specificity | Lesion | SCOPE vs PANORAMA | vs Normal | McNemar | **p<0.001** | **p<0.001** |
| Sensitivity @ 95% Specificity | PDAC | SCOPE vs Baseline | vs Normal | McNemar | **p<0.001** | **p<0.001** |

Classified as Microsoft Confidential

| | | | | | | |
|---|---|---|---|---|---|---|
| Sensitivity @ 95% Specificity | PDAC | SCOPE vs Baseline | vs All | McNemar | p=0.072 | **p<0.001** |
| Sensitivity @ 95% Specificity | PDAC | SCOPE vs PANORAMA | vs Normal | McNemar | **p<0.001** | **p=0.012** |
| Sensitivity @ 95% Specificity | PDAC | SCOPE vs PANORAMA | vs All | McNemar | p=0.271 | **p=0.015** |
| Sensitivity @ 95% Specificity | PanNET | SCOPE vs Baseline | vs Normal | McNemar | **p=0.041** | **p=0.004** |
| Sensitivity @ 95% Specificity | PanNET | SCOPE vs Baseline | vs All | McNemar | **p=0.042** | **p=0.040** |
| Sensitivity @ 95% Specificity | Cyst | SCOPE vs Baseline | vs Normal | McNemar | p=0.748 | p=0.140 |
| Sensitivity @ 95% Specificity | Cyst | SCOPE vs Baseline | vs All | McNemar | p=0.500 | **p=0.041** |

*Table S5: Statistical tests for small-lesion reader study.*

| Reader | Class | Metric | Model | Reader | Superiority P-value | Non-Inferiority P-Value |
|---|---|---|---|---|---|---|
| 1 | PDAC | Sensitivity | 0.775 | 0.8 | 0.7461 | 0.1587 |
| 1 | PDAC | Specificity | 0.9375 | 0.9563 | 0.8666 | **0.0002** |
| 1 | PDAC | PPV | 0.7561 | 0.8205 | 0.8569 | 0.4073 |
| 1 | CYST | Sensitivity | 0.775 | 0.725 | 0.3953 | 0.0544 |
| 1 | CYST | Specificity | 0.9313 | 0.9187 | 0.4159 | **0.0001** |
| 1 | CYST | PPV | 0.7381 | 0.6905 | 0.3089 | **0.0478** |
| 1 | PANNET | Sensitivity | 0.5 | 0.95 | 1 | 0.9902 |
| 1 | PANNET | Specificity | 0.85 | 0.8833 | 0.8659 | **0.0289** |
| 1 | PANNET | PPV | 0.2703 | 0.475 | 0.9894 | 0.893 |
| 1 | LESION | Sensitivity | 0.93 | 0.96 | 0.8867 | **0.0174** |
| 1 | LESION | Specificity | 0.73 | 0.75 | 0.6911 | 0.0912 |
| 1 | LESION | PPV | 0.775 | 0.7934 | 0.8092 | 0.0852 |
| 2 | PDAC | Sensitivity | 0.775 | 0.65 | **0.0898** | **0.0013** |
| 2 | PDAC | Specificity | 0.9375 | 0.9625 | 0.927 | **0.0003** |
| 2 | PDAC | PPV | 0.7561 | 0.8125 | 0.5 | 0.1068 |
| 2 | CYST | Sensitivity | 0.775 | 0.85 | 0.8867 | 0.3815 |
| 2 | CYST | Specificity | 0.9313 | 0.8313 | **0.0057** | **0.001** |
| 2 | CYST | PPV | 0.7381 | 0.5574 | 0.0395 | **0.0014** |
| 2 | PANNET | Sensitivity | 0.5 | 0.95 | 1 | 0.9902 |
| 2 | PANNET | Specificity | 0.85 | 0.9111 | 0.9702 | 0.1371 |
| 2 | PANNET | PPV | 0.2703 | 0.5429 | 0.9987 | 0.9753 |
| 2 | LESION | Sensitivity | 0.93 | 0.98 | 0.9805 | **0.0478** |
| 2 | LESION | Specificity | 0.73 | 0.7 | 0.383 | **0.0263** |
| 2 | LESION | PPV | 0.775 | 0.7656 | 0.6583 | **0.0373** |
| 3 | PDAC | Sensitivity | 0.775 | 0.7 | 0.212 | **0.0163** |

Classified as Microsoft Confidential

| | | | | | | |
|---|---|---|---|---|---|---|
| 3 | PDAC | Specificity | 0.9375 | 0.8875 | **0.0577** | **0.001** |
| 3 | PDAC | PPV | 0.7561 | 0.6 | **0.0288** | **0.001** |
| 3 | CYST | Sensitivity | 0.775 | 0.5 | **0.0133** | **0.0005** |
| 3 | CYST | Specificity | 0.9313 | 0.925 | 0.5 | **0.0002** |
| 3 | CYST | PPV | 0.7381 | 0.625 | **0.0481** | **0.0035** |
| 3 | PANNET | Sensitivity | 0.5 | 0.8 | 0.9961 | 0.9214 |
| 3 | PANNET | Specificity | 0.85 | 0.8778 | 0.8256 | **0.0212** |
| 3 | PANNET | PPV | 0.2703 | 0.4211 | 0.9573 | 0.7536 |
| 3 | LESION | Sensitivity | 0.93 | 0.93 | 0.623 | **0.0008** |
| 3 | LESION | Specificity | 0.73 | 0.78 | 0.8316 | 0.2117 |
| 3 | LESION | PPV | 0.775 | 0.8087 | 0.8042 | **0.0944** |


Classified as Microsoft Confidential
